# Wastewater sequencing reveals the genomic landscape of Influenza A virus in Switzerland

**DOI:** 10.1101/2025.04.17.25325990

**Authors:** Anika John, Seju Kang, Lara Fuhrmann, Ivan Topolsky, Christopher Kent, Joshua Quick, Tanja Stadler, Timothy R. Julian, Niko Beerenwinkel

## Abstract

Influenza A virus poses significant public health challenges, causing seasonal outbreaks and pandemics. Its rapid evolution motivates continuous monitoring of circulating influenza genomes to inform vaccine and antiviral development. Wastewater-based surveillance offers an unbiased, cost-effective approach for genomic surveillance. We developed a novel tiling amplicon primer panel that covers diversity of influenza A virus, targeting segments of the surface proteins HA, NA, and M of subtypes H1N1 and H3N2. Using this panel, we sequenced nucleic acid extracts from 59 Swiss wastewater samples collected at four locations during the 2022/2023 and 2023/2024 winter seasons. We found that wastewater-based abundance estimates of the dominant H1N1 clades correlated with clinical-based estimates in the 2023/2024 season. Furthermore, wastewater-based sequencing revealed mutations in vaccine and drug target sites, consistent with clinical data. We demonstrate the effectiveness of wastewater-based genomic surveillance of influenza A, including lineage identification and mutation tracking to inform vaccine and antiviral strategies.

## Introduction

Influenza A virus poses a substantial burden on the medical and public health systems as it contributes to both seasonal outbreaks and, in severe instances, pandemics^1–4^. The continued public health risk from influenza A is caused in large part by the virus’s high genomic diversity^5^, which facilitates immune escape. The segmented genome structure, consisting of eight negative-sense RNA segments, is a main contributor to its high genomic diversity. An increased chance of genomic reassortment (i.e., genetic exchange)^6,7^ leads to genetic drift and antigenic shift^8^. Hence, influenza A spreads and evolves on a seasonal basis, underpinning the emergence of new strains from mutations that enable the virus to evade immune defenses.

Influenza A antigenicity is determined by the surface glycoproteins hemagglutinin (HA) and neuraminidase (NA) and used for classification of subtypes^9^. The most prevalent influenza A subtypes that cause human infection include (H1N1)pdm09 (hereafter referred to as H1N1), which replaced the previous seasonal H1N1 subtype in 2009^9^, and H3N2 (CDC, 2024). Subtypes of influenza A also circulate in animals^10^ and can spillover into humans, including swine influenza (H1N1, H1N2) and avian influenza (H5N1, H7N9), with a current outbreak of highly pathogenic avian influenza H5N1 spilling over into cattle and people in the United States (CDC, 2025).

Tracking the evolution of influenza A enables detection and identification of circulating subtype strains in the community and can guide the development of effective vaccines. Further, monitoring the genetic diversity of influenza A is a continuous public health effort as mutations in the HA and NA segment of influenza A have been shown to induce low vaccine effectiveness and reduce the efficacy of antiviral drugs ^11,12^.

Wastewater-based surveillance (WBS) has been successfully implemented to monitor spread of SARS-CoV-2 at the community level by quantifying the amount of virus in the wastewater using PCR methods that are cost-effective, unbiased, and granular ^13–15^. WBS has also been used as a platform to monitor the genomic landscape of SARS-CoV-2. Longitudinal sample analysis provided insights into shifts in relative abundance of SARS-CoV-2 variants, as well as emergence of new variants into communities^13^. The emerging SARS-CoV-2 variants of concern were detected in wastewater samples at the University of California San Diego campus up to 14 days earlier than clinical cases^16^. Wastewater sequencing from municipal utility districts in the San Francisco Bay Area revealed the emergence of similar SARS-CoV-2 variants in advance to local California patient-derived genotypes^17^. In addition, more lineages of SARS-CoV-2 were identified in wastewater sequencing data than in the clinical-derived data^18^. In most cases, SARS-CoV-2 genomic sequencing from wastewater has relied on tiling amplicon sequencing using the primer panel designed by the PrimerScheme with demonstrated compatibility across multiple sequencing platforms^13,19^.

Influenza A has also been quantified in wastewater with monitoring of trends in viral concentrations and loads aligning well with the reported clinical data^20,21^. Further, genomic surveillance of influenza A in wastewater has been demonstrated. The relative abundance of influenza A compared to other viruses in wastewater was obtained by sequencing using a viral capture panel^22^. The vaccine-resistant influenza A H3N2, 3C.2a1b.2a.2 subclade, was identified from wastewater sequencing data in Las Vegas, United States^23^. Whole-genome sequencing of influenza A in wastewater has also been demonstrated in Northern Ireland using multi-segment PCR primers which target a conserved region of all eight segments and diverse human and avian influenza A subtypes were identified^24^.

Despite the current success in wastewater sequencing of influenza A, in comparison to SARS-CoV-2, the sequencing coverage from wastewater is low^24^, which may be due to the high diversity of the influenza genome, leading to limited insights into the genomic landscape of circulating influenza A in the community. To provide clinically-relevant insights including relative abundances estimates of circulating clades or identifying emergence and spread of mutations, wastewater sequencing approaches for influenza A need to increase sequencing depth coverage.

In this study, we developed and applied a nucleic acid concentration and extraction protocol, coupled to a tiling amplicon primer panel that targets three influenza A segments HA, NA, and M based on reference genomes of the H1N1 and H3N2 subtypes. These three influenza segments were chosen because HA and NA segments provide insights for designing vaccines and have implications for antiviral drug efficacy. The M segment was also included due to the ongoing development of M-based universal vaccines^25^. With in-depth genomic data of influenza A from wastewater, we implemented a bioinformatics pipeline for data analysis, integration, and interpretation that provides insights into influenza A evolution and hence potential immune escape at the community level.

## Results

### Swiss wastewater-based surveillance programme

Sequencing of influenza A was conducted within the scope of an ongoing national WBS program in Switzerland^26^ surveilling for SARS-CoV-2, influenza A and B and respiratory syncytial virus (RSV) (see Methods). Notably, higher influenza A viral loads were observed in wastewater during the winter season (November to February) at all four investigated wastewater treatment plants (WWTPs), coincident with the seasonality of reported cases of influenza A in Switzerland^28^. A subset of wastewater extracts were selected over the last two winter seasons for sequencing (n = 24 for 2022/2023 and n = 35 for 2023/2024, highlighted in **Fig. 1A**). The concentration of influenza A in the selected wastewater samples are provided in **Supplementary Table S1**.

**Fig. 1.**
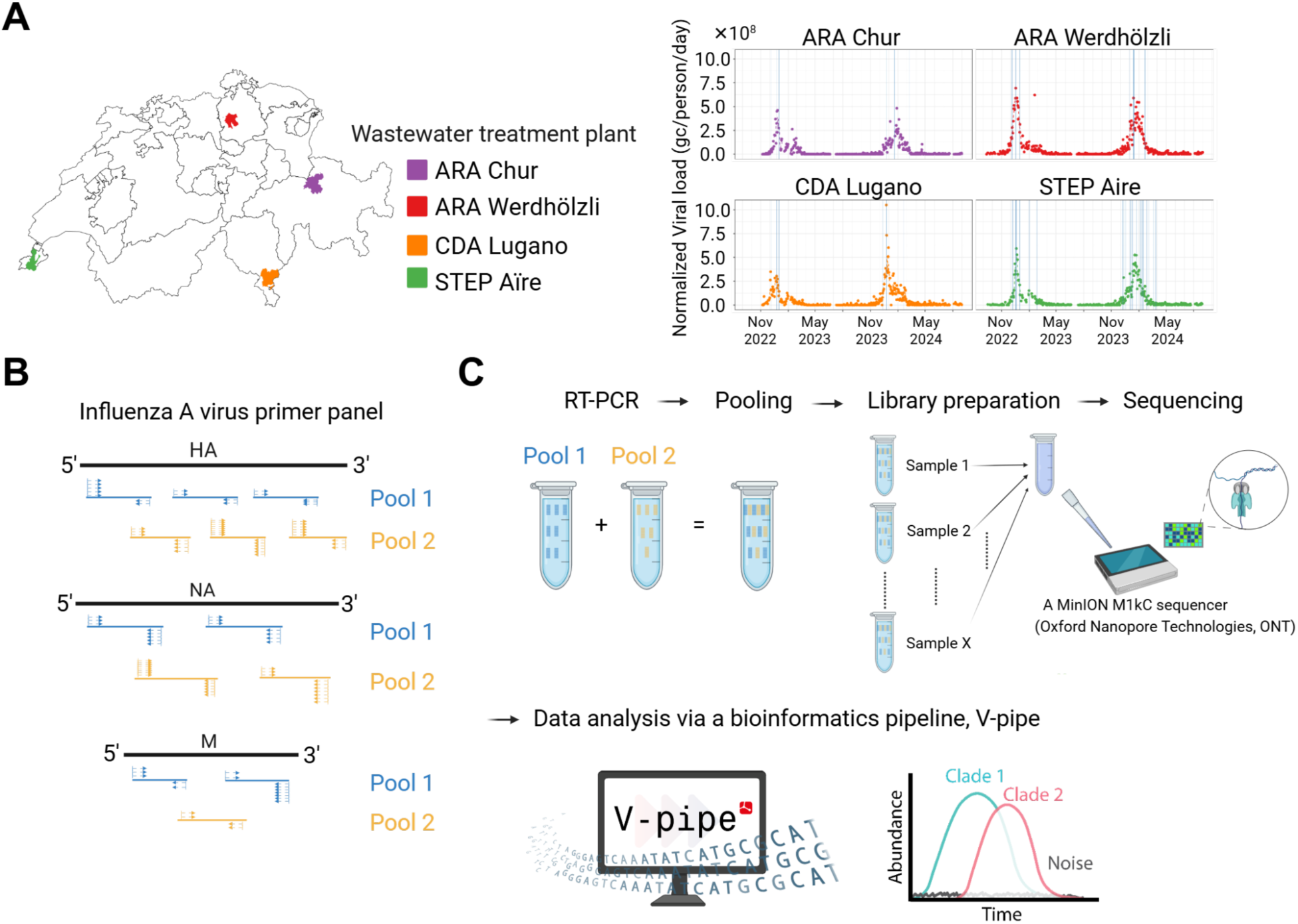
Approach to genomic wastewater-based surveillance of influenza A virus (A) Longitudinal influenza A seasonal dynamics in wastewater collected from four wastewater treatment plants (ARA Chur, ARA Werdhölzli, CDA Lugano, and STEP Aïre) in Switzerland over the last two winter seasons (2022/2023 and 2023/2024). The viral loads in wastewater were estimated using digital PCR assay and normalized by the daily flow rate and resident population served by each of the treatment plants. Blue vertical lines indicate the sample dates selected for influenza A sequencing (B-C) Workflow of tiling amplicon sequencing for influenza A from wastewater extracts (B) Schematic illustration of the primer panel (Pool 1, blue lines; Pool 2, yellow lines) that targets three influenza A segments, HA, NA, and M (black lines). Blue and yellow arrows represent the forward/reverse primers for Pool 1 and 2 (C) Two RT-PCR products from total nucleic acid extracts are pooled and undergo library preparation for sequencing on a MinION M1kC sequencer (Oxford Nanopore Technologies, UK). Sequencing data is analyzed via a bioinformatics pipeline, V-pipe. An example output are clade abundance estimates (light-blue and pink curve). Created in BioRender. Kang, S. (2025) https://BioRender.com/s9rjsbh

### Tiling amplicon sequencing

The primer panel for tiling amplicon sequencing was developed using the latest version of PrimerScheme (v.3.0)^29^ following the ARTIC network protocol^30^. The primer panel consists of two primer mixes (Pool 1 and 2) with a total of 102 primers that generate six amplicons to cover the HA segments, four amplicons to cover the NA segments, and three amplicons to cover the M segments of influenza A H1N1 and H3N2 subtypes. The amplicons are situated to slightly overlap with the neighboring amplicons (**Fig. 1B**). The separate pools contain the primers that target odd (Pool 1) and even (Pool 2) numbered amplicons. A two-step RT-PCR reaction was conducted on the wastewater extracts; complementary DNA (cDNA) synthesis by reverse transcriptases with multi-segment PCR primers and subsequent PCR by the primer panel. The RT-PCR products from both pools were combined for sequencing on a MinION (Oxford Nanopore Technologies, UK). Sequencing data was then analyzed using a customized bioinformatics pipeline integrated into V-pipe^31^ (**Fig. 1C**).

### Quality and validation of influenza A primer panel

Electronic gel electrophoresis of RT-PCR products following amplification of primer Pools 1 and 2 identified clear bands at ∼400 bps, the expected size of the amplicons, demonstrating successful generation of at least one amplicon in the reactions (**Supplementary Fig. S1, Fig. S2**). Further, validating the primer panel on clinical isolates of H1N1 and H3N2 subtypes showed complete coverage with a high read depth (∼10^5^ reads) across all three targets, HA, NA, and M, for both subtypes (**Supplementary Fig. S3**).

To assess the quality of sequencing data derived from wastewater samples, per-amplicon read count, the number of reads covering an amplicon of interest, for HA, NA, and M segments of both subtypes was assessed (**Fig. 2A**). Higher amplicon read counts of HA and NA for H1N1 subtype, referred to as H1 and N1, were obtained from 2023/2024 season than from 2022/2023, which aligns with the clinical surveillance data showing a higher prevalence of H1N1 subtypes in 2023/2024 season (WHO, 2024). Compared to H1 and N1, lower read counts of HA and NA for the H3N2 subtype, here referred to as H3 and N2, were found across most of the amplicons. Notably, however, H3 read counts were higher than H1 counts for amplicon 1 for both seasons, and H3 read counts were higher than H1 counts for amplicons 5 and 6 in 2022/2023 (**Fig 2**).

**Fig. 2.**
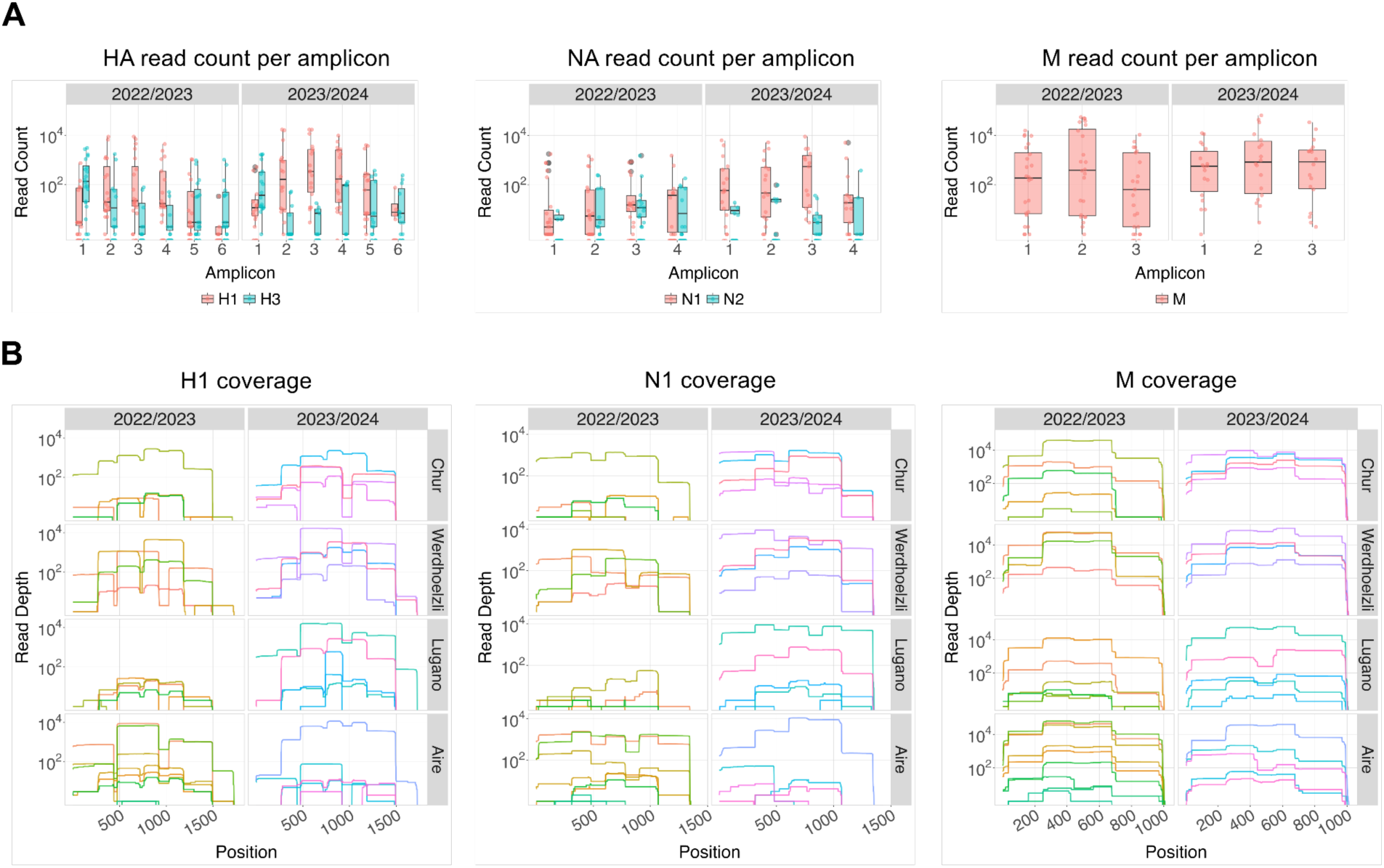
Amplicon read count and position-wise read depth of HA, NA, and M segments obtained from tiling amplicon sequencing of wastewater samples collected from four wastewater treatment plants in Switzerland during the 2022/2023 and 2023/2024 winter seasons. (A) Per amplicon read count distribution as box-whisker plot. The box represents the interquartile range, with the bold line inside indicating the median. The whiskers extend to points within 1.5 times the interquartile range. Values outside this range are displayed as outliers (gray points). The transparent coloured points represent the individual samples. HA and NA corresponding to the H1N1 reference (CY121682.1) are referred to as H1 and N1, and as H3 and N2 for the H3N2 reference (OL693922.1 & KJ609208.1 respectively). (B) Position-wise read depth of H1, N1, and M per influenza A season and wastewater treatment plant. The different colors represent individual samples.

Next, we investigated the position-wise read depth, the number of sequencing reads that align to a specific position in a reference genome, for HA, NA, and M segments of both subtypes (**Fig. 2B** & **Supplementary Fig. S4**). To assess the factors influencing the read depth of a given influenza A target segment, we modeled the mean positional read depth using a zero-inflated negative binomial regression, which allows to account for sequencing drop-outs (see Methods). Based on this model we found a significantly higher mean read depth of the M-segment compared to the other segments (p-value < 2 × 10^−16^, Wald-test). In contrast, the read depth of H3N2 HA-and NA-segment is significantly lower (p-value = 1.7 × 10^−3^, Wald-test). Interestingly concentration as measured using dPCR does not have a statistically significant impact on the mean positional read depth (p-value = 5.2 × 10^−1^, Wald-test)(**Supplementary Table S2**).

### H1N1 clade abundance estimation from wastewater samples

We generated a time-series dataset for the 2023/2024 influenza A season, which was dominated by the H1N1 subtype in Europe^32^. We analyzed weekly wastewater samples between December 2023 and March 2024 (calendar week 49, 2023 to calendar week 12, 2024) from one wastewater treatment (STEP Aïre, Geneva, Switzerland), one of the largest catchment areas in Switzerland. We selected samples up to calendar week eight, as later samples experienced coverage drop-outs likely related to low viral concentrations in wastewater towards the end of the influenza A season (**Supplementary Fig. S5**)^34^. Using this time-series data, we estimated the relative abundance of the two dominating H1N1 clades, 6B.1A.5a.2a and 6B.1A.5a.2a.1 by deconvoluting signature mutation frequencies into clade frequencies over time using the V-pipe integrated tool LolliPop^31,33^ (see Methods). To compare findings to clinical data, weekly clinical abundance estimates were obtained from 9333 H1-segment sequences from across Europe reported to GISAID^34^ (see Methods). We decided to focus on Europe-wide data as data from Switzerland was limited, with only around 250 H1-segment sequences unevenly distributed throughout the same period, which resulted in Swiss clinical data being more noisy and hence less reliable for abundance estimation.

Wastewater-based clade abundance estimates were similar to clinical-based estimates. Specifically, both datasets showed 6B.1A.5a.2a increasing while 6B.1A.5a.2a.1 decreased. However, wastewater-based 6B.1A.5a.2a.1 abundance estimates were slightly higher between week one and four, within the span of the confidence interval (**Fig. 3B & Supplementary Fig. 6**). In addition, for the wastewater estimates, an average 2% of reads covering the variant-defining region could not be assigned to either clade, likely due to sequencing errors.

**Fig. 3.**
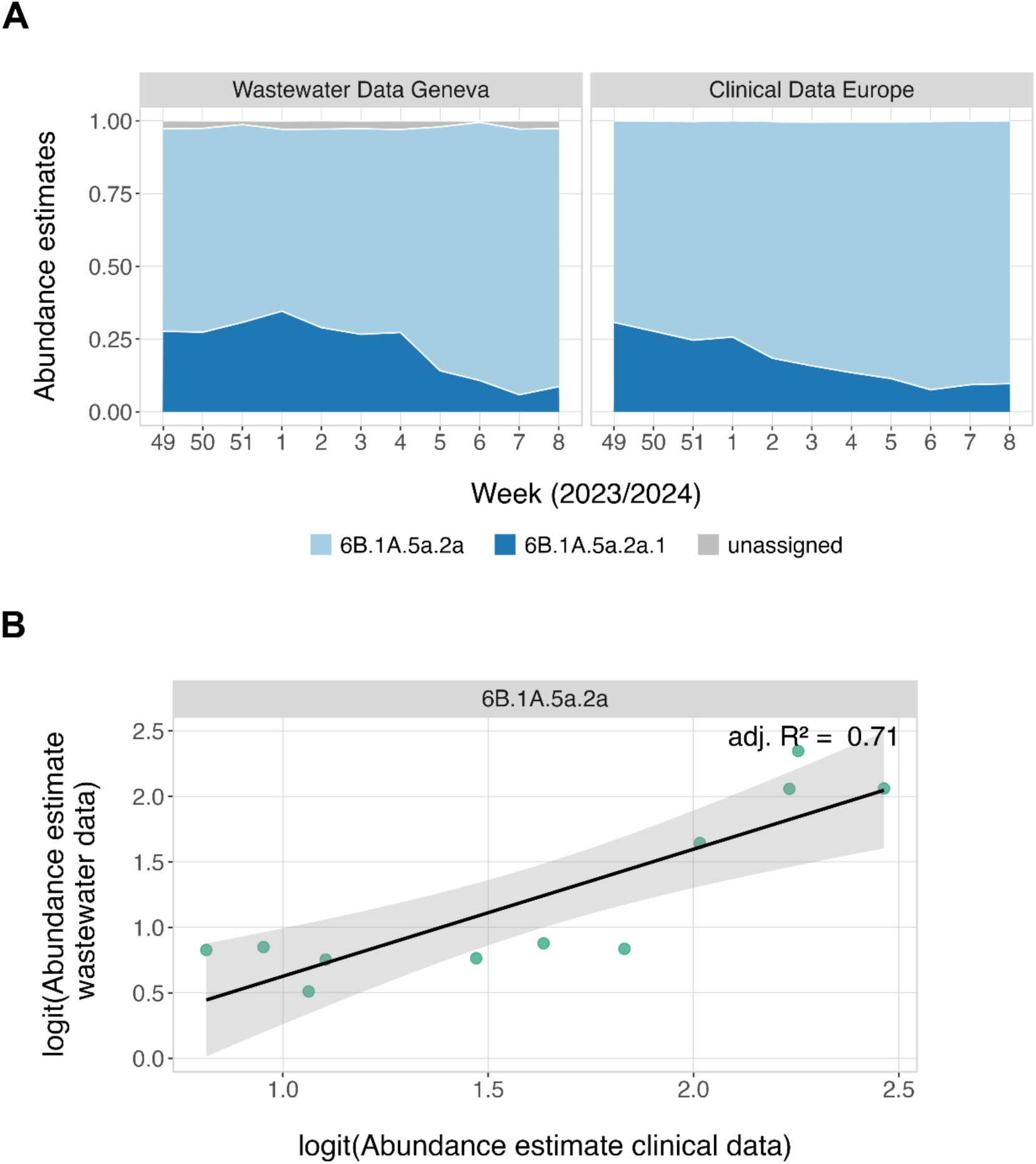
H1N1 subclade abundance estimation from wastewater obtained from Geneva, Switzerland, compared to clinical data from all of Europe. For wastewater data, the relative abundances of 6B.1A.5a.2a and 6B.1A.5a.2a.1 were estimated using LolliPop^33^. Wastewater samples with read depth < 5x at the signature mutations were excluded. The clinical data originated from GISAID^34^ and comprised 9333 samples (for sequence IDs see Source Data 2). (A) Stacked area graph comparing the wastewater and clinical abundance estimates of the dominating H1N1 clades. (B) Comparison of clinical- and wastewater-based 6B.1A.5a.2a abundance estimates on logit scale. The black line represents the derived linear model, the shaded area the 95% confidence interval.

For a quantitative comparison we fitted a simple linear model to logit-transformed wastewater and clinical relative abundance estimates for each clade. The read depth of wastewater samples does not appear to influence the concordance between clinical and wastewater abundance estimates (**Supplementary Figure S7).** For both clades, we observed a statistically significant relationship between wastewater- and clinical relative abundance estimates with slopes close to one, indicating that wastewater estimates are very similar to clinical estimates (6B.1A.5a.2a: R^2^ = 0.71, *β_1_* = 0.97, p-value = 7 × 10^−4^, SE = 0.19, two-sided t-test; 6B.1A.5a.2a.1: R^2^ = 0.66, *β_2_* = 1.03, p-value = 1.56 × 10^−3^, SE = 0.23, two-sided t-test) (**Fig. 3B** and **Supplementary Table S3**).

### Detecting influenza A drug target mutations from wastewater

Alongside annual vaccine recommendations, the WHO aims to contain the public health risk of influenza A through ongoing monitoring of antiviral drug resistance mutations located on the NA segment (WHO 2025). Hence, we investigated the presence of NA mutations of the dominating H1N1 subtype in our wastewater time-series dataset through genomic mutation calling and subsequent conversion of the nucleotide substitution to the respective amino acid substitution (see Methods).

Using the previously described time-series data we detect a single S247N mutation in the late February 2024 sample (calendar week 8, STEP Aïre). This mutation was also observed at low frequencies in clinical samples during the time period of interest, and resulted in mild reduction of the efficacy of the most commonly prescribed antiviral drug, oseltamivir^35^ (**Fig. 4**). The investigated mutations experienced sufficient coverage in the majority of samples (**Supplementary Fig. S8B)**. Our findings indicate that circulating NA mutations, which may have significant implications for public health monitoring, can be robustly detected in wastewater.

**Fig. 4.**
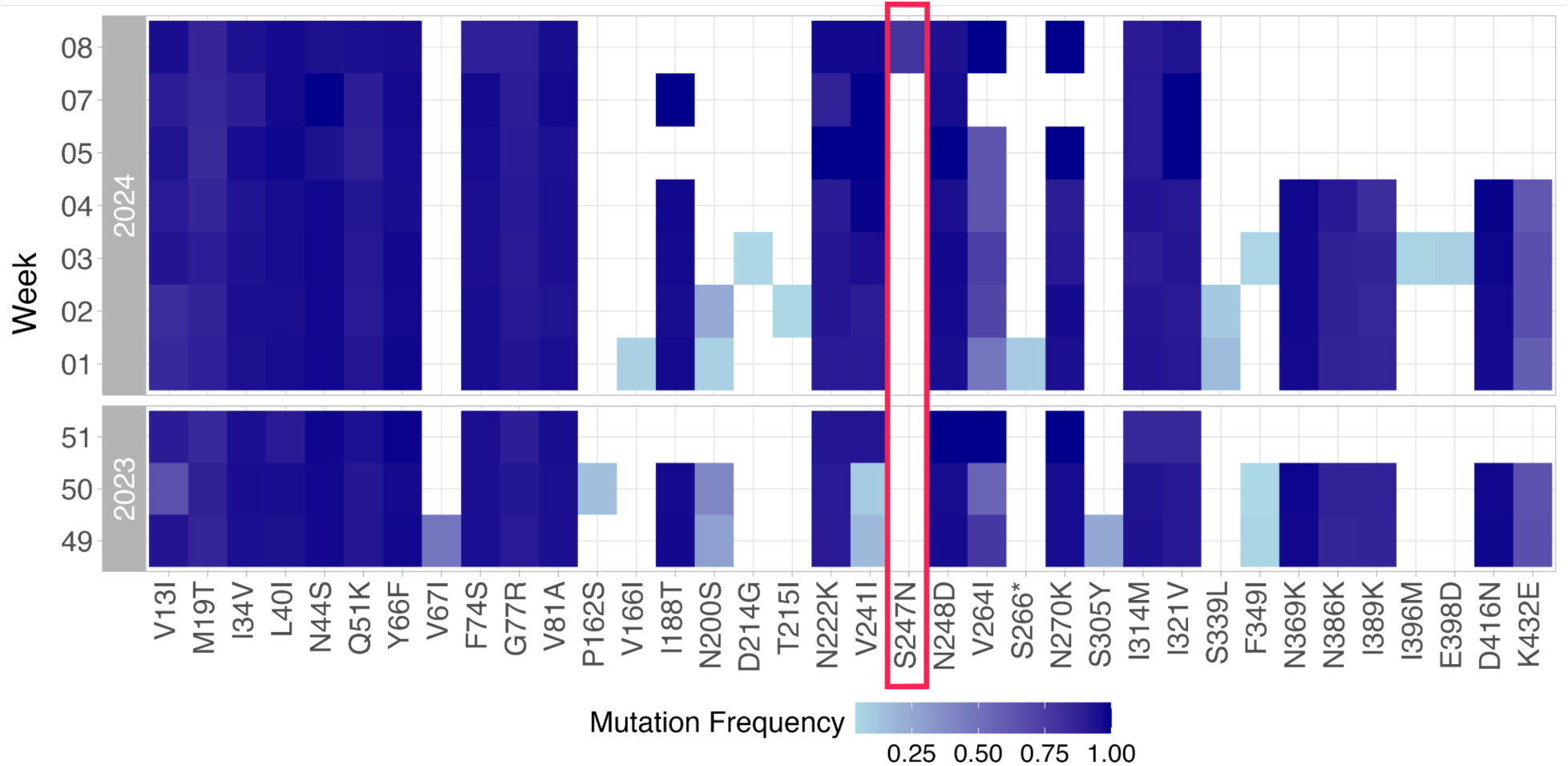
Mutation Frequencies for N1 segment from wastewater samples from one location (STEP Aïre) in Switzerland. The absence of a mutation is marked as white. In red, the known drug resistance mutation S247N is marked. Mutations are relative to the A/California/07/2009 (CY121682.1) reference sequence.

### Monitoring vaccine-relevant mutations using wastewater samples

Using the time-series data, we investigated the genetic diversity when considering vaccine-relevant sites during the influenza A season 2023/2024. Similar to the analysis for N1, we analyzed H1 mutations by calling mutations and converting nucleotide substitutions to corresponding amino acid changes (see Methods). This revealed several low-frequency mutations in HA epitope sites, protein regions critical for antibody binding^36,37^. Notably, we found an increase in frequency over time for K169Q (nucleotide positions 575-577), a signature mutation of the 6B1A.5a.2a clade (WHO, 2024). The majority of epitope site mutations show sufficient coverage in the given samples (**Supplementary Fig. S5B)**.

The influenza matrix protein 2 ectodomain (M2e), encoded on the M segment, is a promising target for universal vaccine design as it contains a large number of conserved epitopes^38^. Using our primer panel, the majority of the M2e domain is located on the same M-segment amplicon (M2e sites 2-9 on amplicon 1, M2e sites 10-24 on amplicon 3). To investigate whether we find the same over-time homogeneity in the M2e domain across our time-series wastewater samples, we reconstructed the viral haplotypes of the M2e domain for each sample. We did so based on separate VILOCA^39^ haplotype reconstructions of the two genomically distant regions of the M2e domain, each spanning 201 nucleotides and covering the positions of interest on amplicon 1 and amplicon 2 respectively (see Methods). We found no difference among M2e haplotypes across wastewater samples, and the resulting consensus sequence is identical to the M2e consensus sequence reconstructed from clinical data collected in Switzerland between December 2023 and March 2024 (**Fig. 5B**). Further, we observe concordance between the wastewater-derived consensus sequence and one M2e consensus sequence (“M2e Literature 1”) presented by Mezhenskaya et al. 2019, which was reported to be closely related to sequences found in swine consistent with the current global circulation of swine origin influenza A(H1N1)pdm09^25^.

**Fig. 5.**
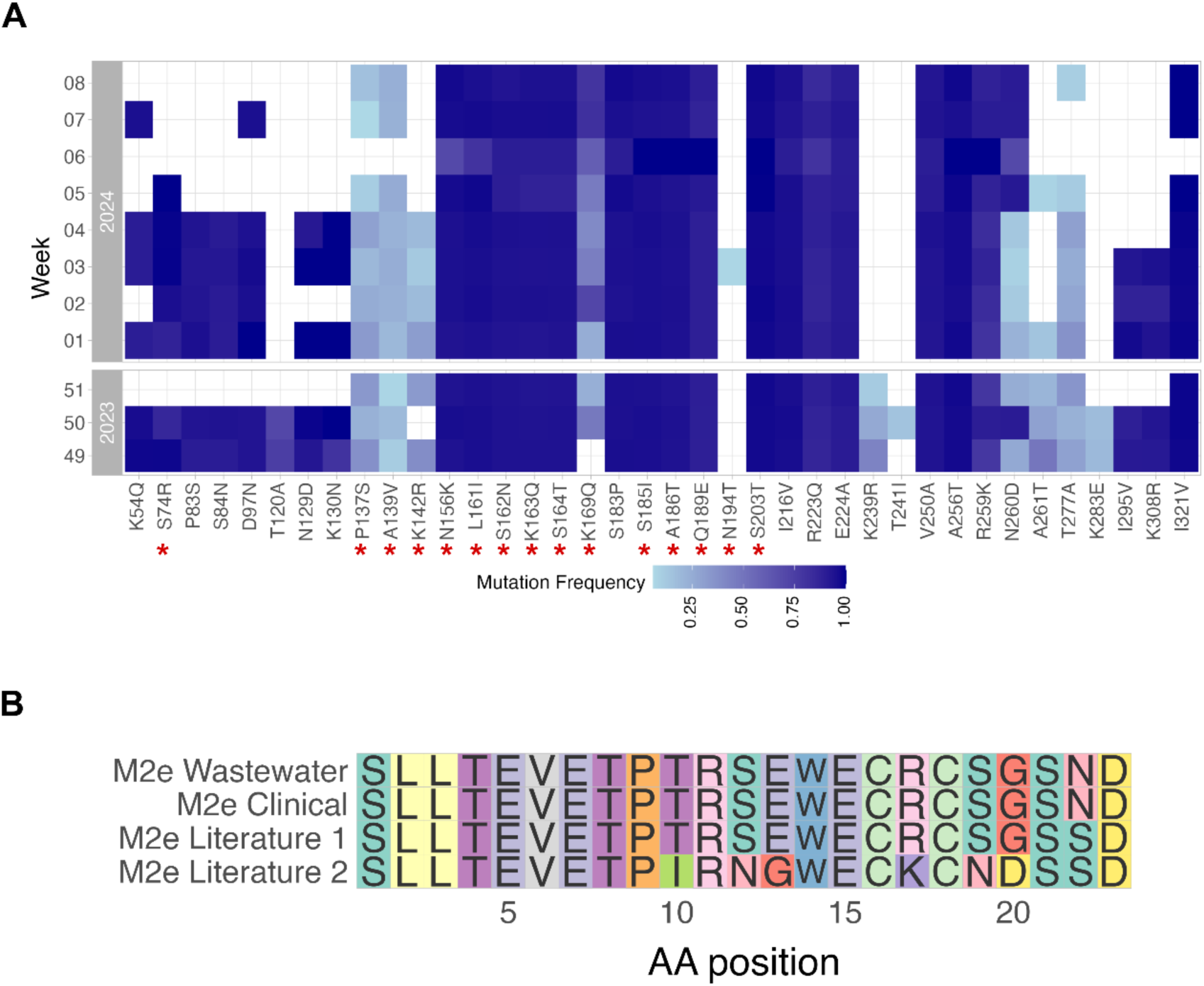
Genomic diversity with respect to vaccine-relevant sites derived from time-resolved wastewater samples (STEP Aïre, Switzerland) (A) H1 mutation frequencies over time relative to reference sequence A/California/07/2009 (CY121680.1) and gene HA0. Red stars mark HA epitope site^37^ (B) Multiple sequence alignment (MSA) of matrix protein 2 ectodomain (M2e) sites on M-segment. M2e wastewater: M2e sequence detected in all wastewater samples of the time-series dataset. M2e Clinical: M2e consensus sequence reconstructed from clinical data collected in Switzerland during December 2023 and March 2024 (GISAID^34^). *M2e Literature 1:* M2e consensus sequence reconstructed by Mezhenskaya et al. 2019 reported to be closely related to sequences found in swine^25^ (lineage 3). *M2e Literature 2:* M2e consensus sequence reconstructed by Mezhenskaya et al. 2019^25^ reported to be closely related to sequences reported prior to the 2009 swine flu pandemic (lineage 5).

Our findings suggest that the genetic diversity of influenza A vaccine-relevant sites observed within wastewater aligns with previous clinical results and that sites, associated with different vaccine types, can be reliably tracked in wastewater.

## Discussion

The continuing evolution of influenza A virus emphasizes the need for genomic surveillance to detect and monitor genetic variants circulating in populations. In this study, we demonstrate genomic sequencing of influenza A virus from wastewater-derived samples based on the development of a novel primer panel for tiling amplicon sequencing and a bespoke bioinformatics pipeline for data analysis and interpretation. Using our methodology, wide coverage with sufficient read depths of three influenza A segments, HA, NA, and M for both H1N1 and H3N2 subtypes was obtained from wastewater samples. Data were collected over the 2022/2023 and 2023/2024 winter seasons at four wastewater treatment plants in Switzerland. The work extends on previous efforts to sequence influenza A by improving substantially on sequencing with high depths required to both detect mutations reliably as well as to estimate the relative abundance of circulating clades. We achieved great sequencing depths through a tiling amplicon approach targeting small amplicons (∼400bp) for the three most epidemiologically relevant segments of the major human prevalent influenza A subtypes. Based on these experimental advances we adapted the existing bioinformatics pipeline V-pipe^31^ for the analysis of wastewater-derived influenza A sequencing data, addressing the challenge posed by the segmented nature of the genome. We developed an approach for influenza A clade abundance prediction from wastewater tiling-amplicon sequences, the first effort in this area. Further we implemented bioinformatics methods to track amino acid changes of interest, both at individual sites and at the level of local haplotypes, which is of particular relevance for vaccine and drug-target sites.

While we have illustrated that genomic sequencing data of influenza A have been successfully acquired from wastewater samples, their quality may be further improved in the future. Typically, sequencing read depth of clinical samples are greater than wastewater ones. For example, using multi-segment PCR primers that bind to termini of influenza A segments^40^, the mean number of reads for HA in clinical samples was 4,764^41^ while it had ∼0 to 191 reads on wastewater samples^24^. Using our primer panel we achieved more in-depth sequencing, with an average ∼836 HA reads across all collected wastewater samples. Despite the wide coverage of three targets, we observed some drop-out regions with relative low read depth. One potential cause to this would be the poor binding of the designed primers to the template (e.g., primer-dimer or secondary structure of the template). Coverage in drop-out regions can be improved by boosting the concentrations of primers or adding additional primers to regions with poor read depth, typically in an iterative manner^42^. Further efforts to improve coverage on drop-outs may be beneficial, particularly if drop-out regions are epidemiologically important, such as being influential in the efficacy of therapeutics or vaccines. The latest version of PrimalScheme used in this study introduces a large number of primers, i.e., primer clouds, on mutative regions without the use of ambiguous bases to handle the diversity for genotypes. More diverse genotypes can be included in the panel to achieve better coverage on drop-outs. An extended discussion on potential enhancements of the primer panel is provided in **Supplementary Discussion**.

Despite possible improvements we have shown that using our primer panel on wastewater samples can provide insightful information about influenza A evolution. Using time-series wastewater samples from the 2023/2024 winter season, we found a statistically significant correlation between clinical- and wastewater-based abundance estimates of 6B.1A.5a.2a and 6B.1A.5a.2a.1, the two dominating H1N1 clades during the 2023/2024 winter season (WHO, 2024). These results confirm the high specificity of our primer panel and demonstrate its effectiveness for clade monitoring in wastewater samples. However, slight deviations in clinical- and wastewater-based abundance estimates were visible, which are not associated with low read depth in wastewater samples. One source of these deviations could be the difference in geographical location. While wastewater samples originated from one location (STEP Aïre) in Switzerland, the clinical data comprised all GISAID-reported H1N1 cases in Europe during the same period. Due to a limitation in the number of available H1N1 sequences from Switzerland we decided against restricting the analysis to Swiss clinical reports. Another reason could be that clinical reports may not fully capture the underlying clade distribution within the population due to sampling biases, whereas wastewater sequencing—as demonstrated for SARS-CoV-2^43^— provides a more unbiased and comprehensive approach.

Wastewater-based sequencing also offers the opportunity to monitor the change of influenza A drug target and vaccine relevant sites over time. In concordance with clinical studies we detected the low frequency N1-mutation S247N in one wastewater sample, which causes mild reduction of oseltamivir inhibition, an antiviral drug frequently recommended by medical organizations^35^ (CDC, 2025; EMA, 2025). Although this mutation emerged towards the end of the influenza A season (WISE: Wastewater-based Infectious Disease Surveillance, 2024), aligning with clinical reports^35^ and providing limited opportunity for observation, the findings show the potential of our method to support WHO’s ongoing NA drug resistance monitoring efforts (WHO, 2025). Further, sequencing HA and M segments from wastewater enabled us to assess the evolution of vaccine-relevant sites. We detected HA epitope site mutations, reported by public health institutions, throughout the 2023/2024 influenza A season. The low-frequency epitope mutations K169Q, P137S and K142R, all located in a high-coverage region, were of particular interest. K169Q, specific to clade 6B1A.5a.2a was first reported by the WHO during this season (WHO, 2024). Although present in an antibody binding site, K169Q was not considered to interfere with vaccine effectiveness and hence was not included in the 2024/2025 vaccine strain^32^ (EMA, 2024). In contrast, P137S and K142R, initially observed during the 2022/2023 season, were included in the 2023/2024 northern hemisphere vaccine strains based on the WHO’s recommendation (WHO, 2023).

Investigating the M2e vaccine target site, present on the M segment, we found no variability in Swiss wastewater or clinical samples during the 2023/2024 season. This aligns with the reportedly high level of conservation in this domain, making it a suitable target for universal vaccines^38,44–46^. In addition, the robustness of our method was further confirmed by the wastewater-derived M2e sequence showing strong similarities to those derived from swine, corresponding to the evolutionary history of H1N1, as swine-flu derived H1N1 strains have been dominant since 2009^47^ (WHO, 2024).

Overall, our findings show that influenza A evolution can be monitored through WBS and inform vaccine and drug development. Due to its unbiased and cost-efficient nature, we believe that this approach has implications for seasonal influenza A monitoring and public health decision making. Indeed, methodologies reported here are implemented for genomic WBS of influenza A in Switzerland ^48^. The continuation of longitudinal monitoring of influenza A genomic information from wastewater is expected to provide robust epidemiological data about influenza A, complementing clinical epidemiology.

## Methods

### Sample collection and processing

The 24h composite raw wastewater influents to four wastewater treatment plants (WWTPs) (CDA Lugano, ARA Werdhölzli, STEP Aïre, CDA Lugano) were collected and transported on ice to the laboratory at Eawag, the Swiss Federal Institute of Aquatic Science and Technology (Dübendorf, Switzerland). Upon arrival, the total nucleic acids were extracted using Wizard® Enviro Total Nucleic Acid Kit (Promega Corporation, Madison, WI, United States; A2991) following the manufacturer’s instructions with minor modifications^21^. In brief, the total nucleic acids were extracted from the 40 mL of composite wastewater influent into 80 µL of RNAse/DNAse-free water. Further, the extract was purified using a OneStep PCR Inhibitor Removal Kit (Zymo Research, Irvine, CA, USA) to remove potential environmental inhibitors. The extracts were stored at −80 °C for up to two years until downstream analysis.

The concentration of influenza A in wastewater was quantified using digital PCR assay, as described in Nadeau et al^21^, and converted to viral load in genome copies per person per day through normalization by the flowrate and resident population for each WWTP^21^.

### Primer panel development

The primer panel was designed based on a representative list of H1N1 and H3N2 target segments (HA, NA and M) obtained from GISAID^34^ for the 2022/2023 season (10.05.2022 – 02.28.2023) in Switzerland. A list of the accession numbers is provided under Source Data 1. To reduce sequence redundancy and avoid bias during primer design, the sequences were aligned using ClustalOmega^49^ and subsequently downsampled using Treemmer (Version 0.3)^50^ with `-RTL 0.95`. From the resulting segment sequences (n = 69 for NA, n = 78 for HA, and n = 82 for M), the influenza A primer panel for tiling amplicon sequencing with an amplicon size of 400 bps was developed using PrimerScheme (v.3.0)^29^ with `--minbasefreq 0`. The panel consists of 102 degenerate primers to account for highly mutated regions, generating six amplicons for HA, four amplicons for NA, and three amplicons for M segments based on an alignment for both subtypes. All primers were synthesized and purchased from Microsynth AG (Balgach, Switzerland). Two primer mixes (Pool 1 and Pool 2) were prepared so that each can amplify odd and even numbered amplicons. The stock concentrations of all primers were adjusted to be 75 nM in the primer mixes with a few 2x concentrated for boosting drop-out regions. The sequences and final concentrations of primers in this panel are provided under Source Data 3 and deposited in the Eawag Research Data Institutional Collection (ERIC) for public availability at https://doi.org/10.25678/000E59.

### Tiling amplicon sequencing

A two-step RT-PCR reaction with this primer panel was conducted on the wastewater extract. The RT step included 2 µL of LunaScript® RT SuperMix (E3010, New England Biolabs (NEB), Ipswich, MA, United States), 2 µL of multi-segment PCR primer sets that bind to termini of influenza A segments^40^ (Tuni 12, 12.4, and 13 (**Supplementary Table S4**) with final concentrations of 160, 40, and 200 nM in the reaction) and 6 µL of wastewater extract. Thermocycling conditions were: 55 °C for 30 s, 42 °C for 50 mins, and 94 °C for 2 mins. Then, cDNA was amplified using the primer panels in the 12.5 µL of reaction volume: 6.25 µL of Q5® High-Fidelity 2× Master Mix (M0492, NEB), 2.5 µL of the primer panel (Pool 1 or 2), and 3.75 µL of cDNA, following the thermocycling conditions: 98 °C for 30 s, 35 cycles of 98 °C for 15 s and 65 °C for 5 mins. After PCR, the products from Pool 1 and 2 were combined for sequencing.

To confirm amplification, electronic gel electrophoresis was conducted on RT-PCR products using Agilent TapeStation 4200 (Agilent Technologies, Santa Clara, CA, United States) with High Sensitivity D5000 ScreenTape Assay (Agilent Technologies; 5067-5592). Then, the RT-PCR products were processed for library preparation for sequencing on the MinION Mk1C sequencer (ONT, Oxford, United Kingdom). Library preparation was conducted using Ligation Sequencing Kit V14 (SQK-LSK114) for a single sample or Native Barcoding Kit V14 (SQK-NBD114.24) for multiple samples. After the library preparation, the sample was loaded onto the R10.4.1 flowcell.

For validation of the primer panel, clinical isolates of two human influenza A subtypes, H1N1 and H3N2, were provided from the Viollier Medical Laboratory (Zurich, Switzerland) and tested along with the wastewater extracts.

### Bioinformatics analysis

The sequencing data in POD5 format was basecalled and demultiplexed (if applicable) into FASTQ format using the Dorado pipeline (Version 7.3.11)^51^ installed on MinION Mk1C. Subsequently, the sequencing reads were processed using the bioinformatics pipeline V-pipe^31^ (Version 3.0) and influenza A subtype- and segment-specific mutation calls, variant abundance estimates and haplotype reconstructions were generated. The pipeline included the main steps of primer trimming using samtools^52^ (Version 1.19), alignment using bwa^53^ (Version 0.7.17), mutation calling using LoFreq^54^ (Version 2.1.3) and, where applicable, local haplotype reconstruction using VILOCA^39^ (Version 1.0.0).

Using the output of the V-pipe alignment step, LolliPop^33^ (Version 0.5.1) was applied to estimate the relative abundance of H1N1 clades, in the December 2023–March 2024 time-series wastewater dataset. To this end 6B.1A.5a.2a and 6B.1A.5a.2a.1 clade-specific mutation (**Supplementary Table S5**) frequencies were deconvolved over time into relative abundance estimates. The kernel bandwidth parameter was set to 1 × 10^−17^. For the clinical abundance estimates, we used all European H1 sequences available on Gisiad^34^ during the same time period; for the accession number list, see Source Data 2. As influenza A clade-classifications of the Gisaid recorded sequences were available in the provided metadata, the weekly frequency of 6B.1A.5a.2a and 6B.1A.5a.2a.1 was calculated by normalizing to the total number of H1 sequences recorded to GISAID that week.

Based on the LoFreq^54^ mutation calling output, nonsynonymous nucleotide point mutations were translated to amino acid mutations using customized scripts (https://github.com/anikajohn/AA_mutationAnalysis_IAV).

The M2e domain, encoded on the M segment and relevant for universal vaccine design, is made up of two genomically distant regions, each covered by one amplicon (M2e nucleotide sites 23-47 on amplicon 1, M2e nucleotide sites 736-780 on amplicon 3). As both regions experienced sufficient read depth in all time-series wastewater samples they were suitable for haplotype reconstruction using VILOCA^39^ with additional parameter settings `--NO- strand_bias_filter --min_windows_coverage 1 --win_coverage 5 -- threshold 0.0` resulting in local haplotypes of length 201 bp (M2e nucleotide sites 23-47 on window “1-201” and nucleotide sites 736-780 on window “671-871”). Subsequently, local haplotypes with a posterior probability below 0.99 and average read number below seven were filtered out. Next nucleotide sequences were converted to amino acid sequences using the Biostrings package in R (Version 2.72.2)^55^ As no variety between sequences of either region existed their consensus sequences were combined to one M2e sequence displayed in Figure 5B.

The data processing and analysis scripts are available on GitHub (https://github.com/cbg-ethz/Influenza_wastewater_analysis).

### Assessing correlation of clinical and wastewater-based clade abundance estimates

To determine whether a significant correlation between the clinical and wastewater-based clade relative abundance estimates exists, we deployed the linear model

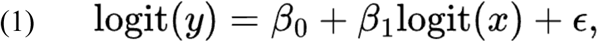

where *y* is the wastewater-based and *x* is the clinical-based relative abundance estimate, *ß_0_* and *ß_1_* are the slope and intercept parameters, respectively, and **ε** is a random error term. The logit-transformation was chosen as it reduced the heteroskedasticity and hence increased the robustness of the model. The model was fitted using the linear regression function implemented in the R package *stats*. The statistical significance of *ß_1_* was assessed using a two-sided t-test. The analysis scripts are available on GitHub (https://github.com/cbg-ethz/Influenza_wastewater_analysis).

### Assessing which factors influence mean position-wise read depth

To evaluate the factors affecting the read depth of a specific influenza A target segment, we modeled the rounded mean positional read depth using a zero-inflated negative binomial regression. The model was fitted using the *glmmTMB()* function of the *glmmTMB* R package^56^ (version 1.1.1). Within the function, the family distribution parameter was set as negative binomial distribution (family = nbinom2) and the zero-inflation equation was set as intercept value (ziformula = ∼ 1). As fixed effects we included the additive effects of concentration (gc/μl), influenza A subtype, experimental batch, wastewater treatment plant and segment type being a nested factor in subtype. As random effects we included additive effects of sample and influenza A season. The statistical significance of the fixed effects were assessed by using the Wald-test, implemented within *glmmTMB*. Model assumptions were tested using the *DHARMa* package (version 0.4.7) in R^57^. The analysis scripts are available on GitHub (https://github.com/cbg-ethz/Influenza_wastewater_analysis).

## Data Availability

Digital PCR data of viral concentration of influenza A virus in wastewater are available for download from https://github.com/EawagPHH/RespiratoryVirusesWastewater and wise.ethz.ch. Wastewater sequencing data are available on the European Nucleotide Archive (ENA) under project accession number PRJEB85534. Clinical sequencing data are available on ENA under sample accession numbers SAMEA117552834 and SAMEA117552704.

## Code Availability

All code used in the analysis and more detailed information about the Bioinformatic analysis is available on https://github.com/cbg-ethz/Influenza_wastewater_analysis.

## Ethical Approval

The Ethikkommission Nordwest- und Zentralschweiz confirmed that the research project does not fall under the scope of the Human Research Act, because the project is not defined as a research project as per HRA Art. 2. An authorisation from the ethics committee is therefore not required.

## Declaration of generative AI and AI-assisted technologies in the writing process

During the preparation of this work the authors used ChatGPT in order to improve the readability of the text. After using this tool/service, the authors reviewed and edited the content as needed and take full responsibility for the content of the publication.

## Acknowledgments

We thank the members of Wastewater Monitoring Laboratory at Eawag for wastewater sample processing including nucleic acid extraction and digital PCR. This study is funded by the Swiss National Science Foundation [Sinergia grant 205933] and the Swiss Federal Office of Public Health (FOPH). S.K. is funded by the Eawag Discretionary Postdoctoral Fellowship. We thank the Viollier Medical Laboratory (Allschwil, Switzerland) for provision of clinical isolates of two influenza A subtypes. We thank Charlyne Bürki and Louis du Plessis for support with the clinical sample selection and logistics. We also thank all collaborators from the Wastewater Infectious Diseases Surveillance and Epidemiology (WISE) project for providing insightful feedback to this project. Further, we gratefully acknowledge all clinical data contributors, i.e., the authors and their originating laboratories responsible for obtaining the clinical specimens, and their submitting laboratories for generating the genetic sequence and metadata and sharing via the GISAID Initiative.

## Contributions

TRJ and NB conceptualized, supervised, and provided funding. AJ and SK designed the study, curated data, performed formal analyses, prepared visualizations, and wrote the first draft of the manuscript. LF provided Methodology support, IT provided Software support, CK and JQ provided technical support on primer panel design. TS is a co-lead of the WISE consortium and provided support on clinical and public health aspects. All authors reviewed, edited, and approved the final version. All authors had full access to all the data in the study and had final responsibility for the decision to submit for publication.

