## Supplementary for "Wastewater sequencing reveals the genomic landscape of Influenza A virus in Switzerland"

### Supplementary Discussion

Coverage was not only low at specific genomic regions, but also for some wastewater samples. We perceive that coverage should be influenced by influenza A concentrations, with lower concentrations leading to lower coverage due to reduced concentrations of target template DNA. However, many wastewater samples had overall poor coverages despite their high influenza A viral concentrations. Interestingly, we found no statistically significant association between influenza A concentration in wastewater and target segment coverage. Yet, HA and NA coverages of both subtypes were significantly lower compared to M coverages, likely caused by higher primer specificity, due to a larger degree of sequence conservation[58]. Although the model allowed to account for sequencing drop-outs through a zero-inflation term, the factors influencing such drop-outs remain unclear and the response variable "mean read depth" is a simplification which cannot fully capture the complexity of coverage patterns.

In addition to coverage optimisation, the primer panel could be further expanded by targeting other segments. For example, the addition of primers targeting the PB1 segment could be a valuable panel expansion, as mutations on this segment have been associated with viral fitness advantages[59]. Notably, the number of primers and concentrations should be carefully evaluated, i.e., primer balancing, to maintain sufficient sequencing depth across the targets.

### Supplementary Figures

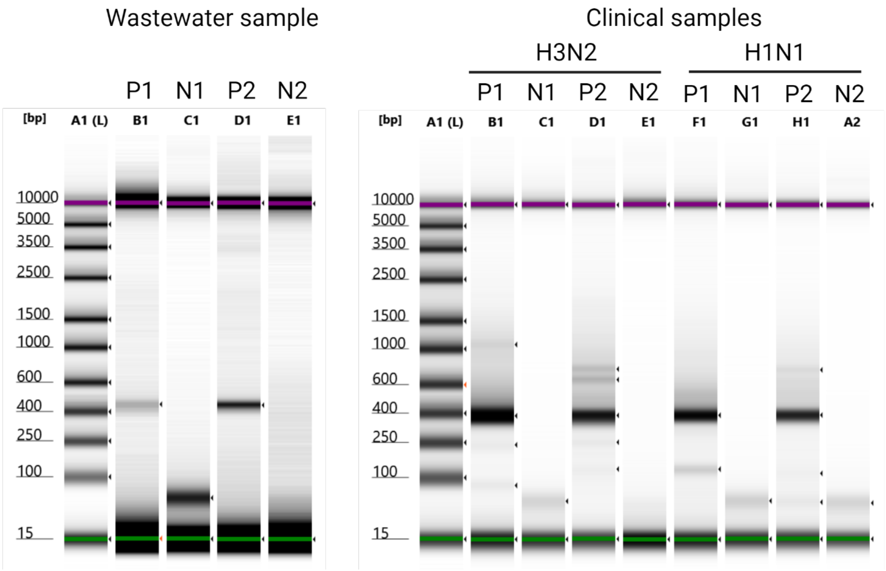

**Fig. S1** Agilent TapeStation 4200 electronic gel electrophoresis of two-step RT-PCR products using the developed primer panel that consists of Pool 1 and 2. The results indicate the samples in the order of reference ladder (A1), samples and no-target control (NTCs) with Pool 1 and 2 (referred to as P1, N1, P2 and N2) on wastewater (left) and H3N2 and H1N1 clinical (right) samples.

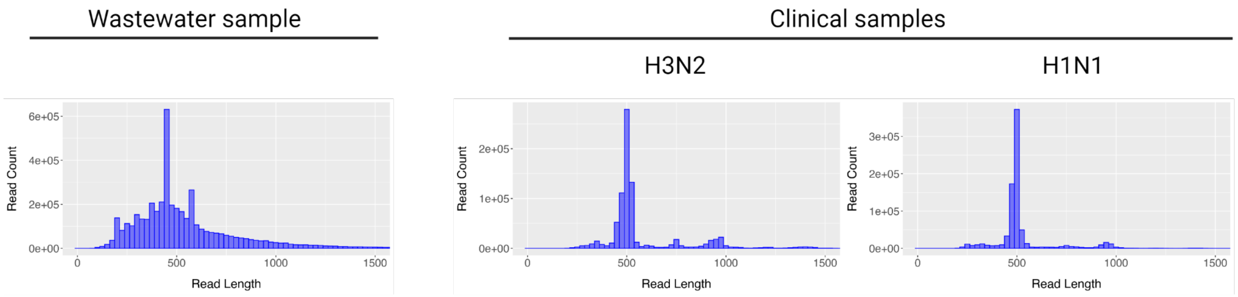

**Fig. S2** Histograms of the number of reads with read lengths in sequencing data. The nucleic acid extracts from wastewater (left) and H3N2 and H1N1 clinical (right) samples were amplified by RT-PCR using two primer mixes, Pool 1 and 2. The pooled samples underwent library preparation and sequenced on a MinION Mk1C sequencer.

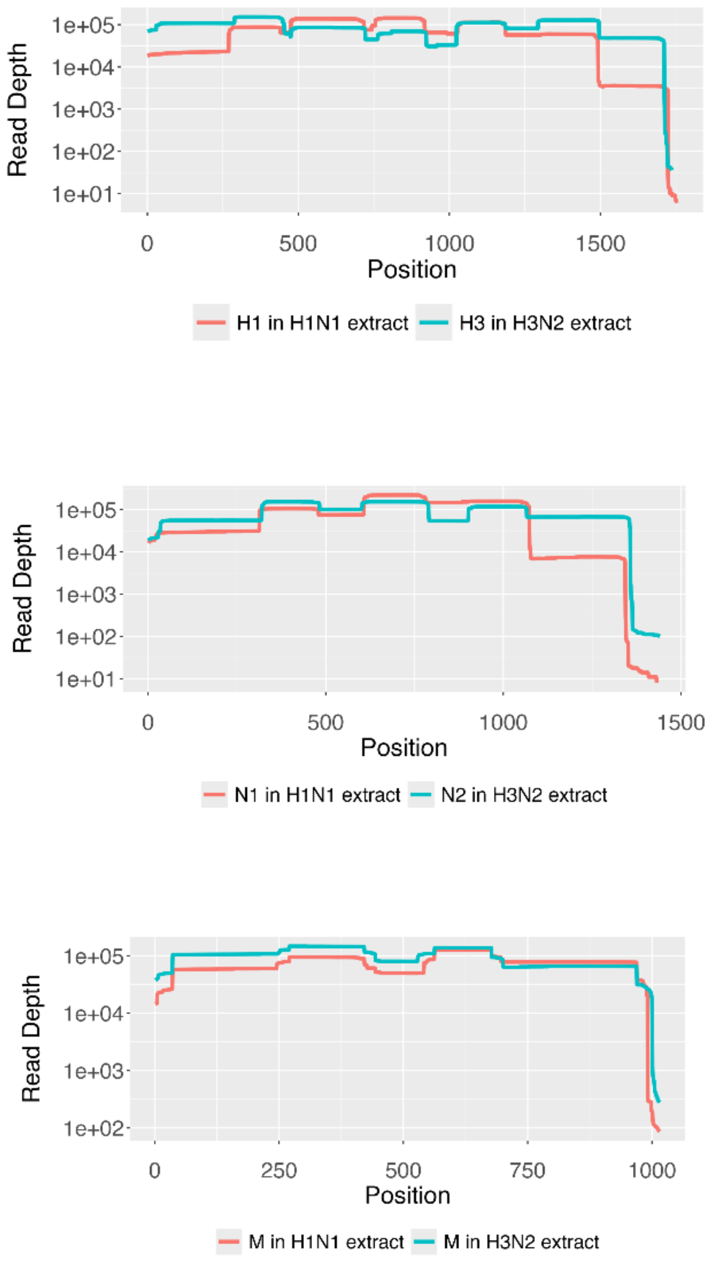

**Fig. S3** Position-wise read depth of HA, NA, and M for H3N2 and H1N1 clinical samples represented by different colors. Sequencing data of H3N2 and H1N1 clinical samples were aligned against the reference of corresponding HA and NA, i.e., H3 (OL693922.1, A/Michigan/05/2021(H3N2)) and N2 (KJ609208.1, A/Perth/16/2009(H3N2)) for H3N2 and H1 (CY121680.1, A/California/07/2009(H1N1)) and N1 (CY121682.1, A/California/07/2009(H1N1)) for H1N1 .

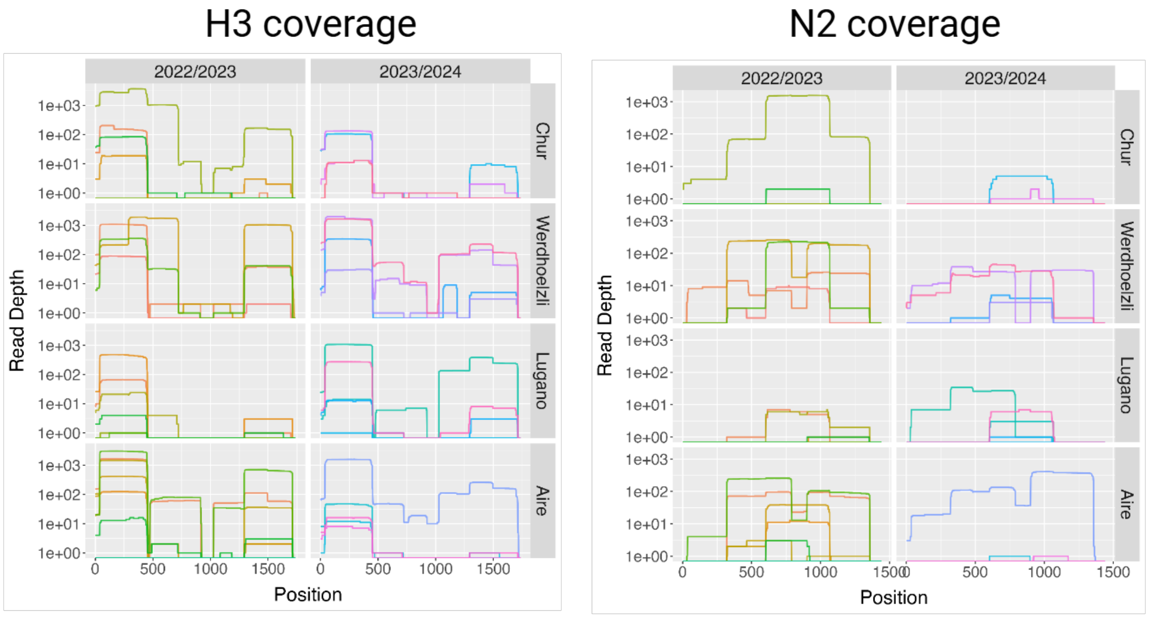

**Fig. S4** Position-wise read depth of H3 (OL693922.1, A/Michigan/05/2021(H3N2)) and N2 (KJ609208.1, A/Perth/16/2009(H3N2)) obtained from tiling amplicon sequencing of wastewater samples collected from four wastewater treatment plants in Switzerland during the 2022/2023 and 2023/2024 winter seasons. The different colors represent individual samples.

**A**

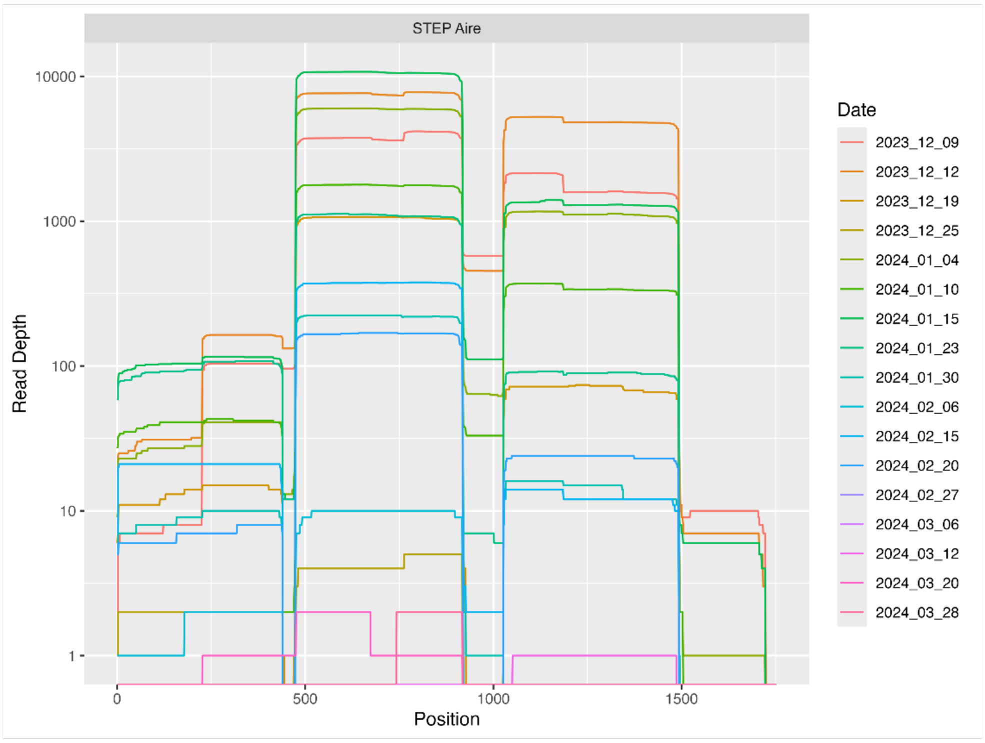
STEP Aire
Read Depth
10000
1000
100
10
1
0
500
1000
1500
Position
Date
2023_12_09
2023_12_12
2023_12_19
2023_12_25
2024_01_04
2024_01_10
2024_01_15
2024_01_23
2024_01_30
2024_02_06
2024_02_15
2024_02_20
2024_02_27
2024_03_06
2024_03_12
2024_03_20
2024_03_28

**B**

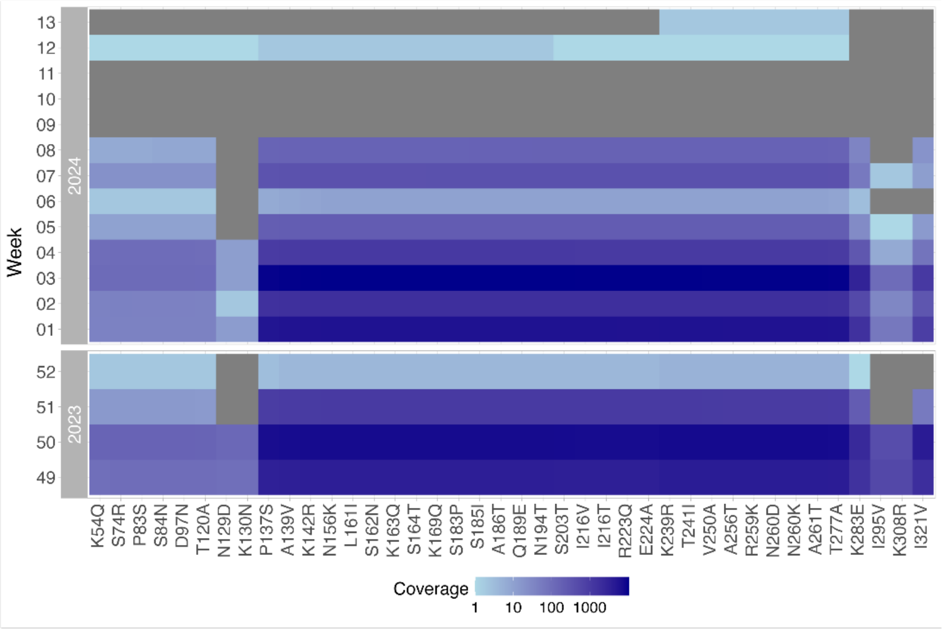
Week
2024
13
12
11
10
09
08
07
06
05
04
03
02
01
2023
52
51
50
49
K54Q
S74R
P83S
S84N
D97N
T120A
N129D
K130N
P137S
A139V
K142R
N156K
L161I
S162N
K163Q
S164T
K169Q
S183P
S185I
A186T
Q189E
N194T
S203T
I216V
I216T
R223Q
E224A
K239R
T241I
V250A
A256T
R259K
N260D
N260K
A261T
T277A
K283E
I295V
K308R
I321V
Coverage
1
10
100
1000

**Fig. S5** Coverage of H1 segment in the time-series dataset (week 49, 2023 to week 12, 2024; (STEP Aïre, Geneva, Switzerland). Alignment was performed relative to the A/California/07/2009 (CY121680.1) reference sequence. (A) Position-wise coverage of H1 segment. The different colors represent individual samples. (B) Coverage of H1 for mutations of interest displayed in Figure 5. Conversion to amino acid mutations was performed relative to gene H0.

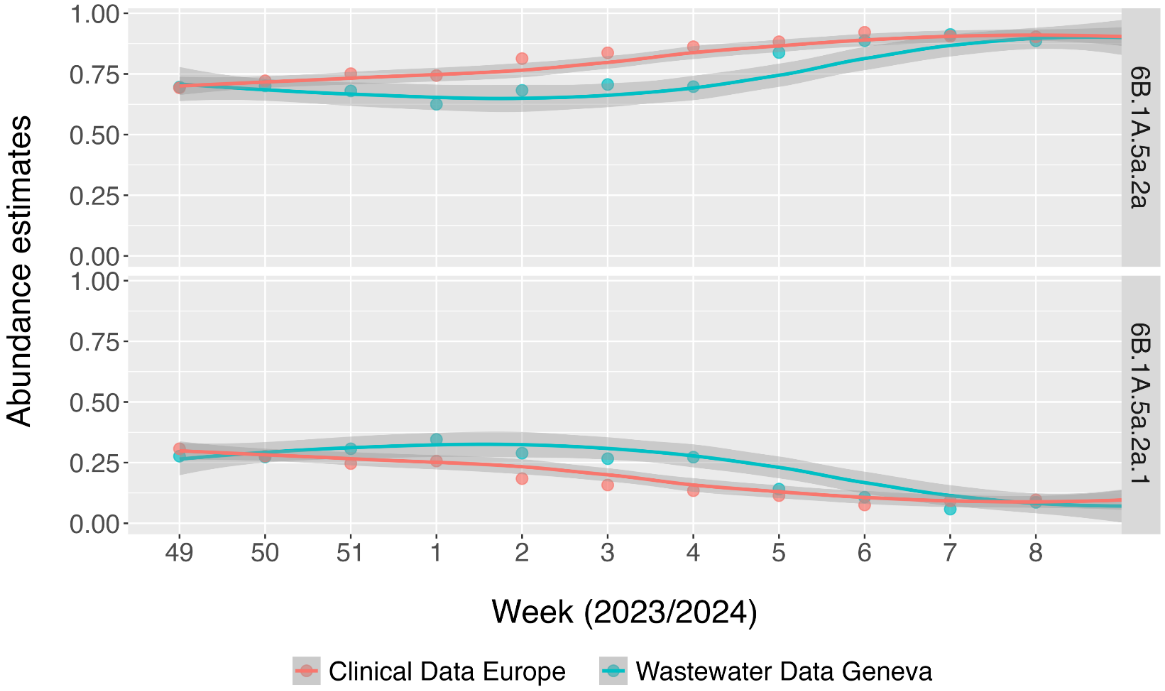

**Fig. S6** H1N1 subclade abundance estimation from wastewater obtained from Geneva, Switzerland, compared to clinical data from all of Europe. The grey-shaded area represents the 95% CI. For wastewater data, the relative abundances of 6B.1A.5a.2a and 6B.1A.5a.2a.1 were estimated using LolliPop[33]. Wastewater samples with read depth < 5x at the signature mutations were excluded. The clinical data originated from GISAID[34] and comprised 9333 samples (for sequence IDs see Source Data 2).

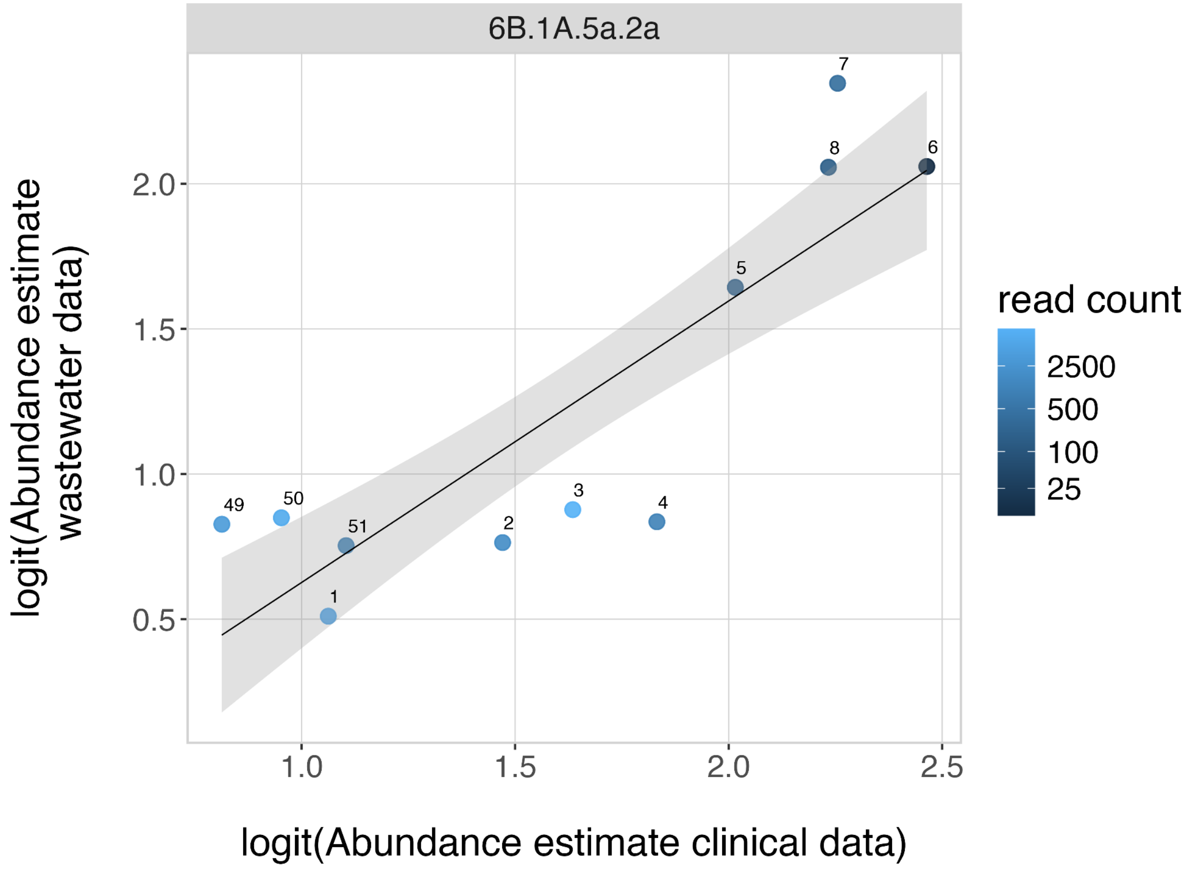

**Fig S7** Comparison of clinical- and wastewater-based 6B.1A.5a.2a abundance estimates on logit scale. The black line represents the derived linear model, the shaded area the 95% confidence interval. Points are coloured by the number of reads covering the variant defining region (on amplicon 3).

**A**

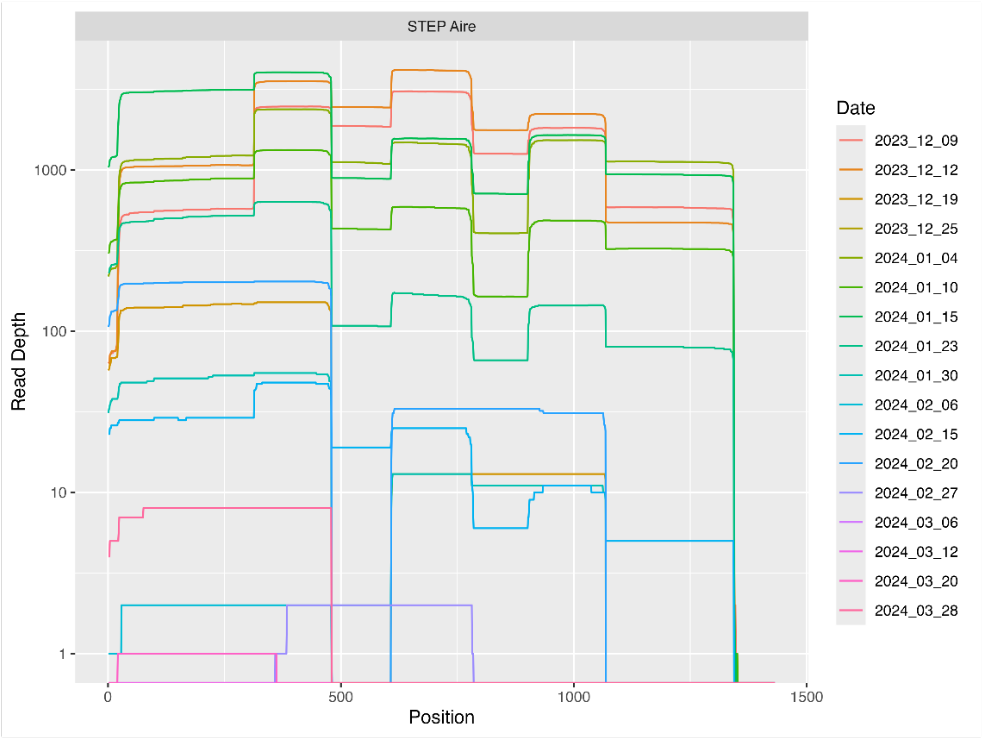
STEP Aire
Read Depth
1000
100
10
1
Position
0
500
1000
1500
Date
2023_12_09
2023_12_12
2023_12_19
2023_12_25
2024_01_04
2024_01_10
2024_01_15
2024_01_23
2024_01_30
2024_02_06
2024_02_15
2024_02_20
2024_02_27
2024_03_06
2024_03_12
2024_03_20
2024_03_28

**B**

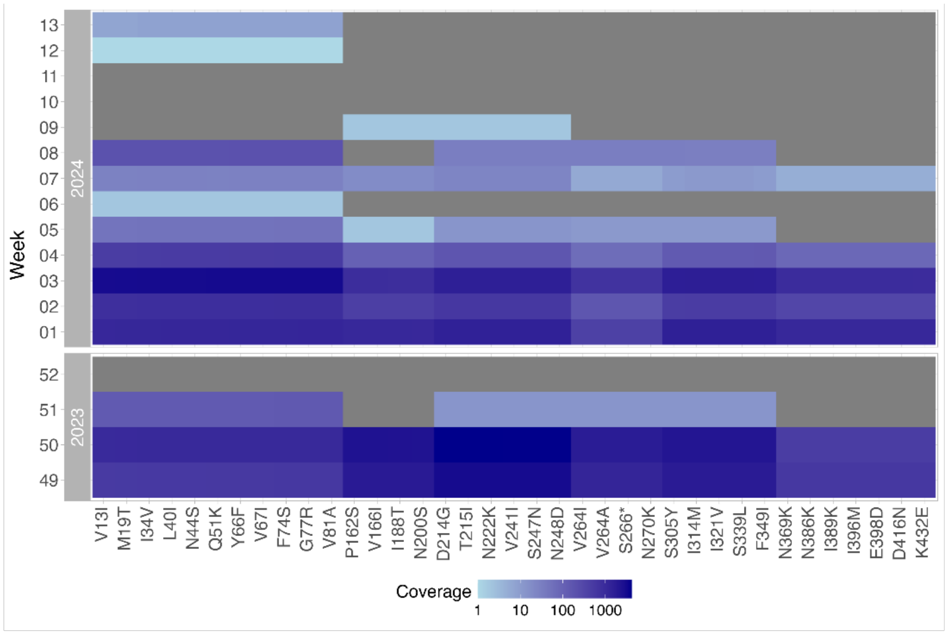
Week
2024
13
12
11
10
09
08
07
06
05
04
03
02
01
2023
52
51
50
49
V13I
M19T
I34V
L40I
N44S
Q51K
Y66F
V67I
F74S
G77R
V81A
P162S
V166I
I188T
N200S
D214G
T215I
N222K
V241I
S247N
N248D
V264I
V264A
S266*
N270K
S305Y
I314M
I321V
S339L
F349I
N369K
N386K
I389K
I396M
E398D
D416N
K432E
Coverage
1
10
100
1000

**Fig. S8** Coverage of N1 segment in the time-series dataset (week 49, 2023 to week 12, 2024; (STEP Aïre, Geneva, Switzerland). Alignment was performed relative to the A/California/07/2009 (CY121682.1) reference sequence. (A) Position-wise coverage of N1 segment. The different colors represent individual samples. (B) Coverage of N1 for mutations of interest as displayed in Figure 4.

### Supplementary Tables

**Table S1** Concentration of influenza A virus in wastewater samples selected in this study measured by digital PCR assay.

| **Season** | **WWTP** | **Date (year_month_day)** | **Influenza A virus concentration in wastewater (gc/mL WW)** | **PCR inhibition on SARS-CoV-2 N1 (-)** | **Mean read-depth of M segment** |
|---|---|---|---|---|---|
| 2022/2023 | CDA Lugano | 2022_12_06 | 37.06 | 0.91 | 216.84 |
| 2022/2023 | CDA Lugano | 2022_12_24 | 88.66 | 0.86 | 14.75 |
| 2022/2023 | CDA Lugano | 2023_01_01 | 46.66 | 0.88 | 4.20 |
| 2022/2023 | ARA Werdhölzli | 2022_12_05 | 52.96 | 0.92 | 200.95 |
| 2022/2023 | ARA Werdhölzli | 2022_12_18 | 226.36 | 0.94 | 22954.20 |
| 2022/2023 | ARA Werdhölzli | 2022_12_31 | 45.46 | 0.89 | 7897.38 |
| 2022/2023 | STEP Aïre | 2022_12_08 | 40.00 | 0.91 | 461.47 |
| 2022/2023 | STEP Aïre | 2022_12_18 | 100.00 | 0.89 | 980.59 |
| 2022/2023 | STEP Aïre | 2023_01_01 | 37.00 | 1.01 | 94.77 |
| 2022/2023 | ARA Chur | 2022_12_10 | 36.26 | 0.93 | 12.24 |
| 2022/2023 | ARA Chur | 2022_12_25 | 203.00 | 0.90 | 17470.48 |
| 2022/2023 | ARA Chur | 2023_01_01 | 102.00 | - | 252.06 |
| 2022/2023 | CDA Lugano | 2022_12_08 | 28.96 | 1.00 | 5477.82 |
| 2022/2023 | CDA Lugano | 2022_12_25 | 77.26 | 0.87 | 2.29 |

| | | | | | |
|---|---|---|---|---|---|
| 2022/2023 | CDA Lugano | 2022_12_31 | 25.60 | 0.93 | 2.69 |
| 2022/2023 | ARA Werdhölzli | 2022_12_06 | 93.00 | 0.93 | 25085.99 |
| 2022/2023 | STEP Aïre | 2022_12_06 | 29.50 | - | 22055.17 |
| 2022/2023 | STEP Aïre | 2022_12_20 | 193.76 | 1.01 | 16264.07 |
| 2022/2023 | STEP Aïre | 2022_12_31 | 59.50 | 1.03 | 31096.64 |
| 2022/2023 | STEP Aïre | 2023_01_28 | 19.22 | 0.95 | 12.56 |
| 2022/2023 | STEP Aïre | 2023_02_02 | 73.02 | - | 3.29 |
| 2022/2023 | STEP Aïre | 2023_02_26 | 17.50 | 0.93 | 1.31 |
| 2022/2023 | ARA Chur | 2022_12_06 | 23.76 | 0.92 | 862.78 |
| 2022/2023 | ARA Chur | 2022_12_29 | 36.76 | - | 1.23 |
| 2023/2024 | CDA Lugano | 2023_12_14 | 37.76 | 0.93 | 16.98 |
| 2023/2024 | CDA Lugano | 2023_12_25 | 214.50 | 1.04 | 49.46 |
| 2023/2024 | CDA Lugano | 2024_02_19 | 366.50 | 1.08 | 1274.37 |
| 2023/2024 | ARA Werdhölzli | 2023_12_31 | 30.76 | 1.13 | 3891.43 |
| 2023/2024 | ARA Werdhölzli | 2024_01_14 | 150.30 | 0.98 | 593.09 |
| 2023/2024 | ARA Werdhölzli | 2024_02_21 | 33.76 | 1.02 | 6418.05 |
| 2023/2024 | STEP Aïre | 2023_12_23 | 41.50 | 1.19 | 25.87 |
| 2023/2024 | STEP Aïre | 2024_01_02 | 58.50 | 1.13 | 18343.60 |
| 2023/2024 | STEP Aïre | 2024_02_12 | 36.76 | 0.91 | 11.29 |
| 2023/2024 | ARA Chur | 2023_12_24 | 39.76 | 1.10 | 2657.03 |
| 2023/2024 | ARA Chur | 2024_01_21 | 248.26 | 1.12 | 438.53 |
| 2023/2024 | ARA Chur | 2024_03_09 | 57.86 | - | 1169.38 |
| 2023/2024 | CDA Lugano | 2023_12_12 | 29.26 | - | 30069.17 |
| 2023/2024 | CDA Lugano | 2023_12_24 | 301.76 | 1.05 | 2.41 |

| | | | | | |
|---|---|---|---|---|---|
| 2023/2024 | ARA Werdhölzli | 2024_01_16 | 89.10 | - | 44988.44 |
| 2023/2024 | STEP Aïre | 2023_12_21 | 31.50 | - | 978.91 |
| 2023/2024 | STEP Aïre | 2024_02_11 | 34.50 | 0.91 | 307.50 |
| 2023/2024 | ARA Chur | 2024_01_20 | 94.26 | 0.97 | 5227.88 |
| 2023/2024 | STEP Aïre | 2023_12_09 | 6.50 | 1.03 | 25636.17 |
| 2023/2024 | STEP Aïre | 2023_12_12 | 23.00 | 1.00 | 33259.35 |
| 2023/2024 | STEP Aïre | 2023_12_19 | 37.76 | 1.11 | 2959.71 |
| 2023/2024 | STEP Aïre | 2023_12_25 | 45.76 | 1.02 | 734.90 |
| 2023/2024 | STEP Aïre | 2024_01_04 | 48.76 | 0.87 | 29122.35 |
| 2023/2024 | STEP Aïre | 2024_01_10 | 57.50 | 1.01 | 26268.12 |
| 2023/2024 | STEP Aïre | 2024_01_15 | 75.26 | 1.01 | 46970.55 |
| 2023/2024 | STEP Aïre | 2024_01_23 | 116.00 | 0.94 | 15602.25 |
| 2023/2024 | STEP Aïre | 2024_01_30 | 102.50 | 0.99 | 3548.30 |
| 2023/2024 | STEP Aïre | 2024_02_06 | 91.32 | 0.89 | 557.40 |
| 2023/2024 | STEP Aïre | 2024_02_15 | 43.50 | 1.02 | 6235.26 |
| 2023/2024 | STEP Aïre | 2024_02_20 | 52.26 | 0.91 | 1644.54 |
| 2023/2024 | STEP Aïre | 2024_02_27 | 16.76 | 1.16 | 1.21 |
| 2023/2024 | STEP Aïre | 2024_03_06 | 9.30 | 0.89 | 13.37 |
| 2023/2024 | STEP Aïre | 2024_03_12 | 19.66 | 0.89 | 0.82 |
| 2023/2024 | STEP Aïre | 2024_03_20 | 9.60 | 0.84 | 0.27 |
| 2023/2024 | STEP Aïre | 2024_03_28 | 8.86 | - | 453.74 |

**Table S2** Table stating zero-inflated negative binomial regression model parameters. Significant levels were determined using a Wald-test.

| | Parameter | Variance | std. deviation | | |
|---|---|---|---|---|---|
| **Additive random effects** | Sample | 7.290 | 2.700 | | |
| | Season | $3.778 \times 10^{-12}$ | $1.944 \times 10^{-6}$ | | |
| **Additive fixed effects** | **Parameter** | **Estimates** | **std. error** | **z value** | **p-value** |
| | Intercept | 3.751 | 1.151 | 3.258 | $1.12 \times 10^{-3}$ |
| | Concentration [gc/µl] | $8.081 \times 10^{-3}$ | $1.260 \times 10^{-2}$ | 0.641 | 0.521 |
| | H3N2 Subtype | -0.808 | 0.258 | -3.137 | $1.70 \times 10^{-3}$ |
| | M both Subtypes | 2.610 | 0.248 | 10.529 | $< 2 \times 10^{-16}$ |
| | Experimental batch | -0.551 | 0.892 | -0.617 | 0.537 |
| | Werdhoelzli | 2.117 | 1.337 | 1.576 | 0.115 |
| | Lugano | -1.364 | 1.248 | -1.093 | 0.275 |
| | Aire | -0.451 | 1.221 | -0.370 | 0.712 |
| **Nested fixed effects** | H1N1:NA | -0.290 | 0.239 | -1.213 | 0.225 |
| | H3N2:NA | -2.372 | 0.286 | -8.295 | $< 2 \times 10^{-16}$ |
| **Zero-inflation model** | Intercept | -21.22 | 3510.17 | $6.0 \times 10^{-3}$ | 0.995 |

**Table S3** Table stating linear model parameters. These parameters are used to display the response curve in Fig. 3B. Significant levels were determined using a two-sided t-test.

| Clade | Coefficient | Estimates | std. error | t-value | p-value |
|---|---|---|---|---|---|
| 6B.1A.5a.2a | Intercept | -0.343 | 0.333 | -1.031 | 0.329 |
| | Slope | 0.970 | 0.194 | 4.993 | $7.46 \times 10^{-4}$ |
| 6B.1A.5a.2a.1 | Intercept | 0.292 | 0.399 | 0.732 | 0.483 |
| | Slope | 1.029 | 0.2303 | 4.466 | $1.56 \times 10^{-3}$ |

**Table S4** Sequence of universal primers that bind to the termini of eight influenza A virus segments.

| Name | Sequence (5'-3') |
|---|---|
| Tuni12 | ACGCGTGATCAGCAAAAGCAGG |
| Tuni12.4 | ACGCGTGATCAGCGAAAGCAGG |
| Tuni13 | ACGCGTGATCAGTAGAAACAAGG |

**Table S5** Clade signature mutations relative to CY121680.1 reference sequence. Signature mutations were determined based on the WHO report *Influenza virus characterization: summary report, Europe, March 2024* (6B.1A.5a.2a.1 mutation corresponds to reported amino acid substitution T277A).

| Clade | Segment | Nucleotide Position | Nucleotide Reference (CY121680.1) | Nucleotide Mutation |
|---|---|---|---|---|
| 6B.1A.5a.2a | HA | 742 | A | C |
| 6B.1A.5a.2a | HA | 847 | G | A |
| 6B.1A.5a.2a | HA | 900 | A | A |
| 6B.1A.5a.2a.1 | HA | 900 | A | G |
